# CHST6-related macular corneal dystrophy: a matter of endothelium

**DOI:** 10.1101/2021.05.31.21258099

**Authors:** Bi Ning Zhang, Benxiang Qi, Xin Wang, Chunxiao Dong, Jun Cheng, Dewei Li, Suxia Li, Min Chen, Bin Zhang, Qingjun Zhou, Lixin Xie

## Abstract

Macular corneal dystrophy (MCD) is classified as corneal stromal dystrophy. In this study, we retrospectively reviewed the surgical outcomes of 118 MCD patients receiving surgical treatment in the past 30 years and found patients receiving penetrating keratoplasty had the lowest recurrence rate 13.75%, compared with 40.91% patients receiving deep anterior lamellar keratoplasty and 25% receiving phototherapeutic keratectomy. Transcriptomic analysis in human corneal single-cell sequencing atlas found the MCD pathogenic gene *CHST6* was abundant in corneal endothelium rather than other cell types. CHST6 protein showed a similar expression pattern to its mRNA. The mouse homologous gene *Chst5* was 120-fold higher in corneal endothelium than in the epithelial and stromal layers. Mice with specifically *Chst5* knockdown in the endothelial layer by microinjection of the adeno-associated virus serotype 9 - shRNA plasmids into the anterior chamber, rather than *Chst5* knockdown into the stroma, showed MCD-like phenotypes. Corneal opacification and abnormally larger collagen fibrils were observed in the endothelial *Chst5* knockdown mice. The same corneal characteristics were observed after overexpressing human *CHST6* mutant R50H in the mouse endothelium. These observations indicating the pathogenesis of MCD is more related to the corneal endothelium rather than the stroma.

**Significance Statement:** Our study gave evidence that corneal endothelium contributing more to the macular corneal dystrophy (MCD) development, rather than other cell types in the cornea. We proposed penetrating keratoplasty might serve as a more proper surgical treatment for MCD according to the recurrence rate analysis. We also provided a novel method to construct MCD mouse model.

## Introduction

Macular corneal dystrophy (MCD, MIM number 217800) involves grey-white irregular opacities in the corneal stroma region and was assigned to stromal dystrophies according to the International Classification of Corneal Dystrophies (IC3D) (1, 2). A progressive corneal clouding generally occurs in both eyes from the first decade of life and will end up with entire vision loss(3). Corneal transplantation is usually required at the middle age of the patient (3). Despite the shortage of cornea tissues available for transplantation (4), there is a high chance MCD patients would suffer recurrence and require regrafting (5).

MCD is inherited in an autosomal recessive manner and the gene *CHST6* is reported to be responsible for the disease (2, 6). At least 181 variants in this gene have been found related to MCD (7). *CHST6* encodes an enzyme corneal *N*-acetylglucosaminyl-6-O-sulfotransferase (corneal GlcNAc6ST; cGn6ST) and functions during keratin sulfate (KS) biosynthesis by transferring sulfate from donor molecule 3’-phosphoadenosine 5’-phosphosulphate (PAPS) to KS glycosaminoglycan (GAG) chains (6, 8). KS constitutes the three corneal proteoglycans: lumican, keratocan, and mimecan, which are essential in maintaining corneal transparency (9).

Besides stroma, Descemet’s membrane lesion was also reported in an MCD patient carrying compound heterozygous *CHST6* mutations (10). Scanning electron microscopy analysis found GAG accumulation in corneal endothelium cytoplasm (11), indicating MCD could affect endothelium as well. Whether endothelium is involved in MCD progression affects the surgical method doctor chooses to treat the disease. Previously we conducted a retrospective study following 51 MCD patients for 18 years and found a lower recurrence rate and delayed recurrence onset in patients undergoing penetrating keratoplasty (PKP) than patients undergoing deep anterior lamellar keratoplasty (DALK) (12). The five-year recurrence rate of PKP is 7.7% compared with 49.5% of the patients undergoing DALK (12). Another study reviewed 84 eyes with lattice corneal dystrophy (LCD) and MCD undergoing PKP or DALK, and found MCD patients with DALK suffered from a progressive decrease in endothelial density, which was not observed in the LCD group. 12 months after DALK, MCD eyes had more than 600 endothelia/mm^2^ less than the LCD eyes, suggesting MCD might not be the optimal corneal dystrophy for DALK treatment (13). A more recent study involving 109 MCD eyes with PKP and 21 MCD eyes with DALK found a higher graft survival rate in PKP group than DALK group, being 93% versus 80% at 1 year time and 78% versus 70% at 4 years, though the graft failure reasons varied (14). Phototherapeutic keratectomy (PTK) is preferred as an early intervention for MCD. However, a study following up 10 MCD eyes undergoing PTK for more than 13 years reported a recurrence in 90% of eyes at less than 4 years post-operation (15). PKP was performed as an amendment in 6 recurrent eyes and none got recurrence during the follow-up period (15). Some groups generated similar views according to the clinical observations, that MCD showed more abnormalities in the posterior cornea (10, 16, 17), and PTK was the more suitable therapy (17). While PKP involves the transplantation of all layers of the cornea, DALK does not involve the transplantation of the corneal endothelium. Based on these findings, we envisioned a scenario that the corneal endothelium is the determinant in this disease. To test this hypothesis, we first evaluated *CHST6* and *Chst5* expression patterns in human and mice cornea respectively. Then we eliminated the gene expression in the endothelial and stromal cells respectively with a location-specific injection of adeno-associated virus serotype 9 (AAV9)-shRNA plasmids and determined the corneal opacification and MCD symptoms by clinical examinations and KS staining.

## Results

### Penetrating keratoplasty contributing to the lowest recurrence rate

A total of 118 MCD patients with 150 eyes who obtained surgical treatment in Qingdao Eye Hospital were included in the study. Recurrence was defined as typical MCD haze extended in the graft button under slit lamp examination regardless of the visual acuity change. As was shown in Table 1, 25% (4/16) of the MCD patients obtaining PTK, 40.91% (9/22) obtaining DALK, and 13.75% (11/80) obtaining PKP were diagnosed as recurrence. The MCD recurrent rate was found associated with the surgical method (*P* = 0.017, χ^2^ test). The average time for recurrence was 7.00 ± 0.82 years, 2.56 ± 2.71 years, and 6.38 ± 3.34 years for PTK, DALK, PKP, respectively (Table 1). PKP showed the lowest recurrence rate among the three types of surgeries. More than half of the PKP recurrent cases (54.5%) were still normal at 5 years post-surgery while 87.5% of the DALK recurrent cases showed MCD signs again in the first four years post-surgery (Fig. 1A). Slit lamp photo of a typical recurrent case after PKP was presented (Fig. 1B). OCT indicating the PKP case was more likely to gain recurrence from the peripheral host cornea (Fig. 1C).

**Table 1.** Recurrent MCD cases with different surgical methods.

| Surgery | Case | Recurrent case | Recurrent case rate | Eye | Recurrent eye | Recurrent eye rate | Average recurrence time (years after first surgery) |
| --- | --- | --- | --- | --- | --- | --- | --- |
| PTK | 16 | 4 | 25% | 26 | 8 | 30.77% | 7 |
| DALK | 22 | 9 | 40.91% | 23 | 9 | 39.13% | 2.56 |
| PKP | 80 | 11 | 13.75% | 101 | 11 | 10.89% | 6.38 |

**Fig. 1.**
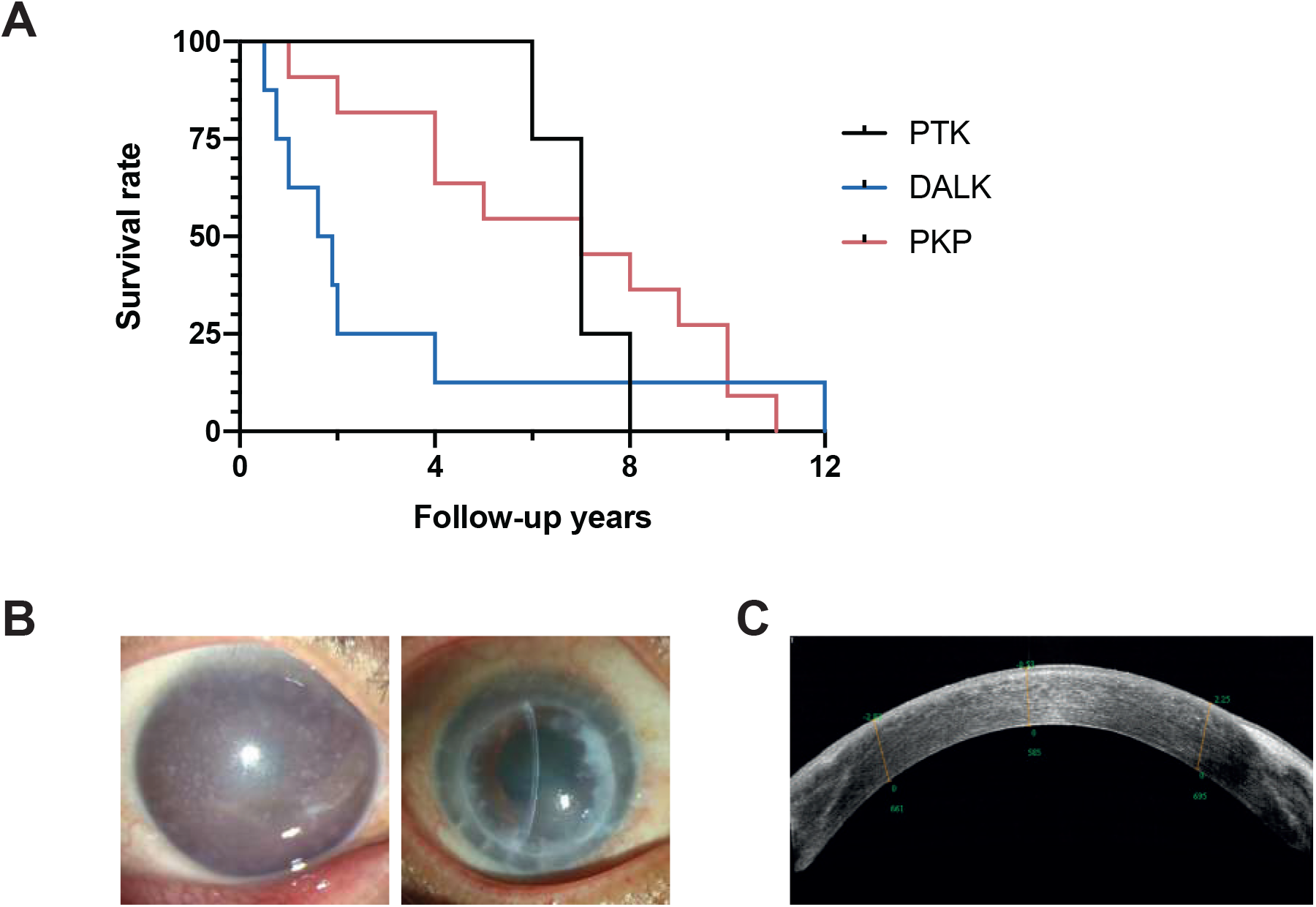
Macular corneal dystrophy recurrent cases. (*A*) Recurrent cases graft survival curve after PTK, DALK, or PKP. A total of 4 PTK recurrent cases, 9 DALK recurrent cases, and 11 PKP recurrent cases were presented. (*B*) Typical recurrent eye after PKP. Slit lamp photos of the right eye of an MCD patient before PKP in 2007 and after recurrence in 2021. Recurrent corneal opacities could be observed at the edges of the graft. (*C*) OCT image of the right eye after the recurrence of the same patient.

### Expression pattern of CHST6 in human cornea

After searching our human corneal single cell sequencing dataset (Fig. 2A), we found *CHST6* was highly expressed in the endothelial cluster rather than in other clusters (Fig. 2B), with the gene detected in around 49.9% of cells in the endothelial cluster and around 2.2% in other cell types. Consistent with the mRNA expression pattern, dark staining of CHST6 protein was present in the endothelium and only sparsely stained CHST6 in the stromal area (Fig. 2C). There were also condensed signals in the basal layer of the epithelium (Fig. 2C). In contrast, MCD cornea gave little expression of CHST6 protein product (Fig. 2D), suggesting the patient carrying a loss-of-function mutation in CHST6. In all, CHST6 is enriched in the human corneal endothelium at both mRNA and protein levels rather than in the stroma.

**Fig. 2.**
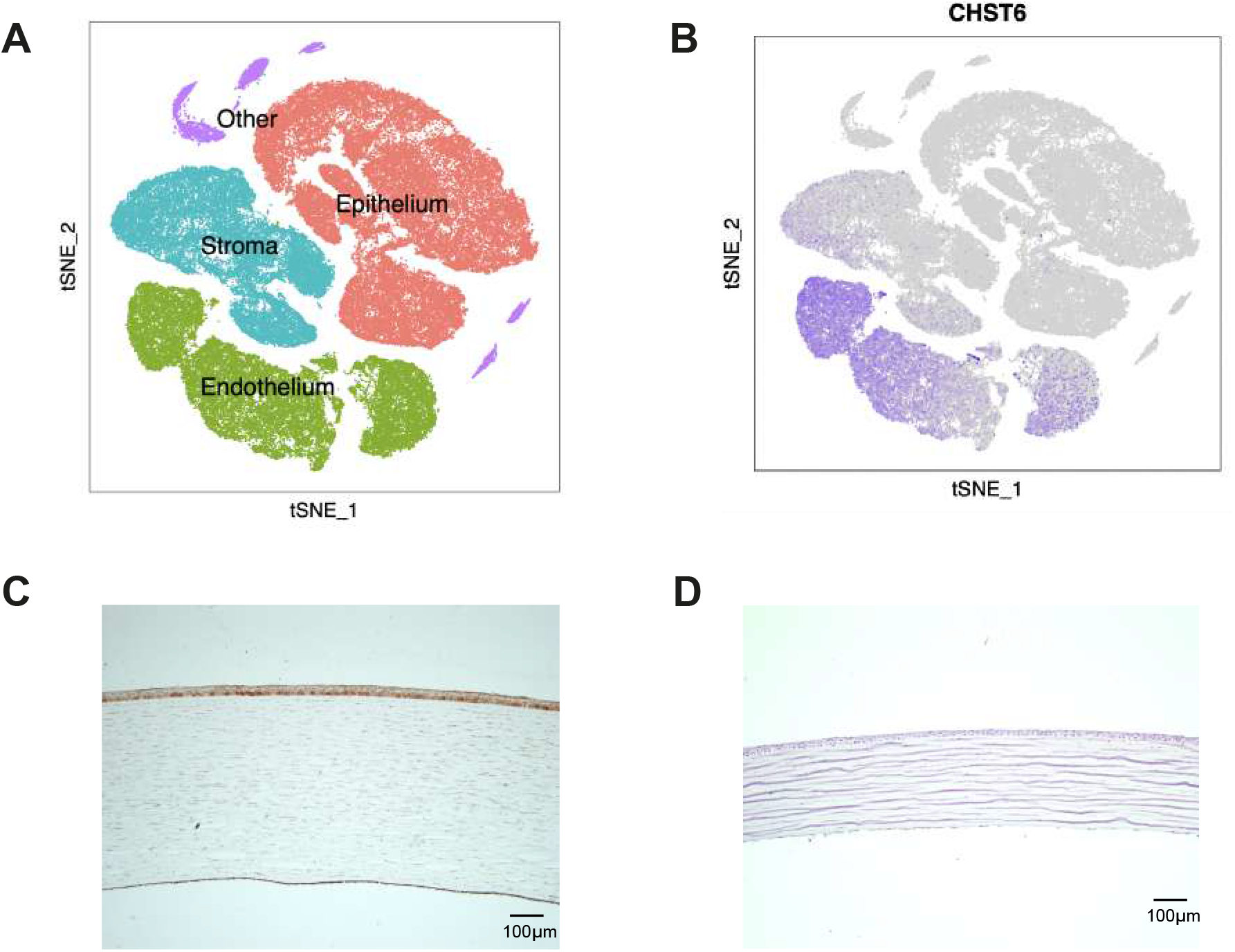
*CHST6* is enriched in corneal endothelium. (*A*) Single-cell transcriptomic atlas of the human cornea. Four clusters were separated by unbiased t-SNE clustering and cell types were annotated. (*B*) t-SNE visualization of *CHST6* in the whole corneal atlas. *CHST6* was positively marked in most of the endothelial cluster and a small number of stromal cells. (*C*) Immunohistology staining of CHST6 in normal human cornea. Dark brown staining could be detected in the endothelial layer and the basal epithelia. Stroma contained sparsely distributed light brown staining. (*D*) Immunohistology staining of CHST6 in MCD patient cornea. No brown staining could be detected in all layers of the diseased cornea.

### Knockdown of endothelial Chst5 contributing to mice corneal opacification

*Chst5* is the mouse homolog of human *CHST6* and the gene showed an expression pattern resemble *CHST6* in human cornea, that the endothelial *Chst5* was more than 120-fold of that in other corneal layers (n=30, *P* < 0.01, unpaired t-test) (Fig. 3A). We adopted an intracameral microinjection method (AH) to knockdown the endothelial *Chst5* (Fig. 3B). Injection of AAV9-shRNA into the anterior chamber could effectively knockdown the endothelial *Chst5* (Fig. 3C). A control shRNA sequence targeting no gene was packaged to the same AAV9 vector serving as the control plasmid. The expression of endothelial *Chst5* in the AH knockdown (AH KD) mice was around 36% of the AH control (AH Con) mice. When adjusted to the other parts of the cornea (epithelium+stroma), the efficacy increased to knockdown 70% more endothelial *Chst5* in AH KD than in AH Con (n=30, *P* = 0.015, unpaired t-test). During intracameral injection, the needle went through the corneal stroma as well. To exclude the possibility that the phenotype observed was caused by the leakage of shRNA into the stroma, we constructed an intrastromal control model by directly injecting the AAV9-shRNA into the corneal stroma (Str) (Fig. 3B), which displayed effective knockdown of *Chst5* in the stroma (Str KD), but not in the endothelium (Fig. 3C). The endothelial *Chst5* was 8.26-fold higher in Str KD than AH KD group while the stromal *Chst5* in Str KD was only 15% of AH KD group. When adjusted endothelial *Chst5* expression with other corneal layers, Str KD showed the highest while AH KD showed the lowest relative *Chst5* expression (Fig. 3C).

**Fig. 3.**
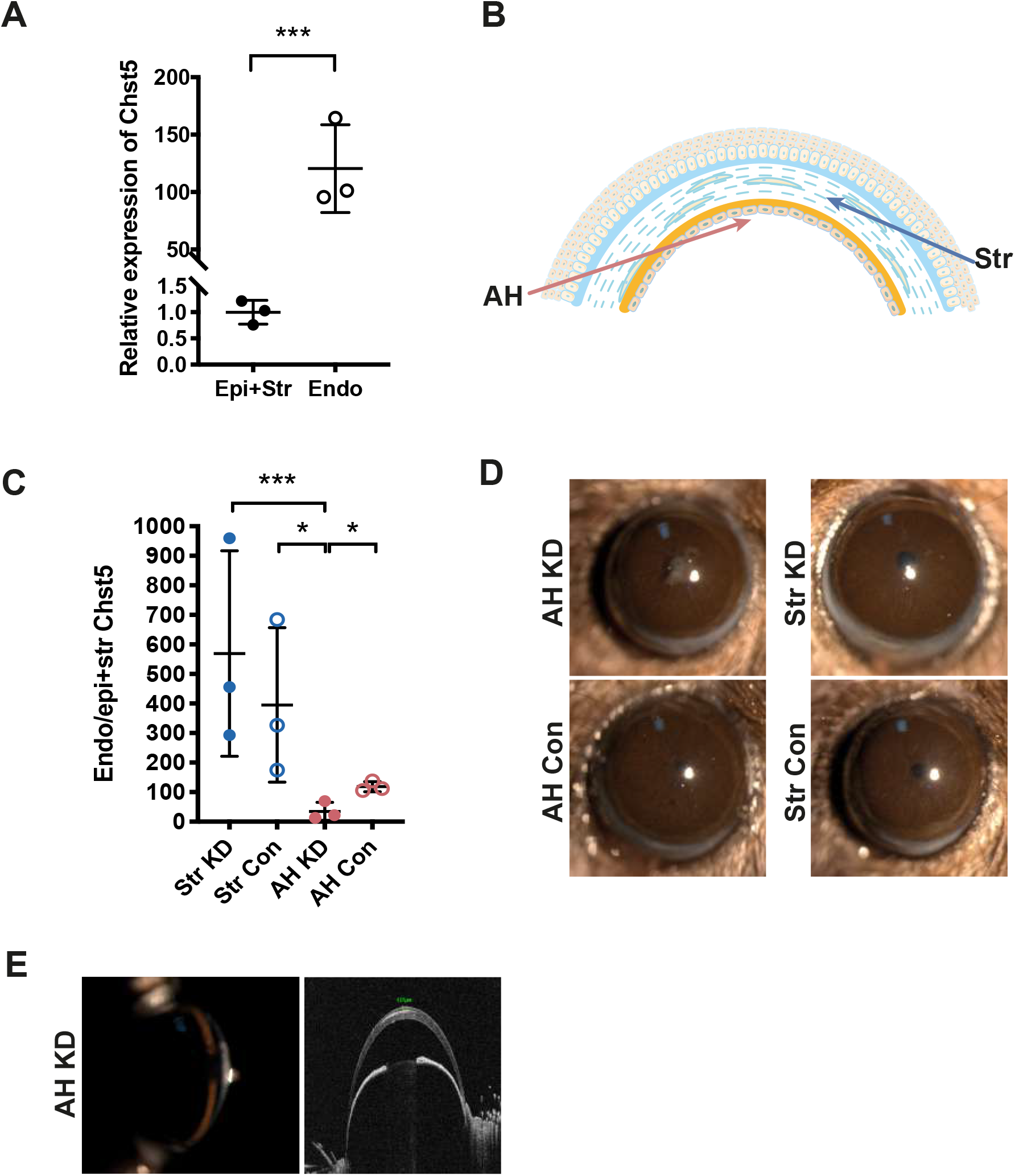
Endothelial *Chst5* was related to corneal opacification. For (*A*) and (*B*), each dot contains 10 mice and a total of 30 mice were included in each group. (*A*) The relative expression level of *Chst5* in endothelial layer and epithelial + stromal layers of normal mice. (*B*) Schematic diagram of two ways of AAV9-shRNA plasmids injection. Pink: intracameral injection route; Blue: intrastromal injection route. (*C*) The fold change of endothelial *Chst5* relative to epithelial and stromal *Chst5* in four treatment groups. A nonparametric Mann-Whitney U test was applied due to the large standard deviation in Str groups. *P* values equal to 0.077, 0.017 between AH KD and Str KD, AH KD and Str Con, respectively. *P* = 0.015 between AH KD and AH Con with unpaired t-test. (*D*) Broad-beam illumination slit lamp photos of a representative mouse in each treatment group. Only AH KD mice showed corneal opacification while corneas in the other three groups kept transparency. (*E*) Slit lamp and anterior segment OCT photos of the (*D*) AH KD mouse. White deposits could be observed in the full thickness of the cornea. Hyper-reflective area could be observed in the central stroma using OCT horizontal line scanning.

Consistent with our hypothesis that MCD corneal haze was related with endothelial Chst5, there were 11.3% (6/53) mice developed MCD-like opacities in the AH KD group, whereas none of the AH Con (0/32), Str KD (0/14), Str Con (0/9) mice developed such opacity (Fig. 3D). The opacity diffused to the full thickness of the corneal stroma (Fig. 3E), similar to that observed in the advanced MCD patients.

### Cornea characteristics after endothelial Chst5 knockdown

KS is the major GAG in the cornea and highly sulphated KS is essential to corneal transparency. cGn6ST encoded by *CHST6* mediates KS sulphation and sulphated KS is detectable by the monoclonal antibody (18). We stained the KS in our Chst5 knockdown mice. Sulphated KS signal was barely detectable at the opaque area in the AH KD mouse while homogenous staining could be seen in Str KD mice cornea (Fig 4A). Alcian blue is an amphoteric dye reacting with acid mucopolysaccharide and has been used as a classic method to detect the accumulation of acid mucopolysaccharide in MCD stroma (19). Consistent with the antibody staining, AH knockdown cornea but not the Str knockdown cornea showed alcian blue positive staining clusters (Fig 4B). Neither of the control AAV9 plasmids displayed abnormal GAG deposits (Fig 4B). As an alteration of acid mucopolysaccharide content could occur in keratopathy with calcification, we examined the same mice corneas with von Kossa staining for calcified deposits as well. No von Kossa positive staining was identified in the alcian blue positive cornea (Fig 4C). A calcific band keratopathy slide was set as the positive control for the staining (Fig 4C). Finally, we applied transmission electron microscopy (TEM) to characterize the structural changes in the cornea. TEM recognized collagen fibrils with larger diameters with dark staining in the AH KD mouse compared with other groups (Fig 4E), which was also presented in the stroma of MCD patient (Fig 4D). This type of abnormal large collagen fibril was also observed by another group in the posterior stroma of MCD patients (16), indicating our AH KD mouse shared similar corneal stromal characteristics with MCD patients. The average collagen fibril diameter was 32.22 ± 2.13 nm in normal mouse, 34.47 ± 2.28 nm in Str KD mouse, and 39.23 ± 3.69 nm in AH KD mouse, respectively (Fig 4F). Though no corneal opacification was observed in the Str KD group, there were alterations in the stromal collagen fibrils.

**Fig. 4.**
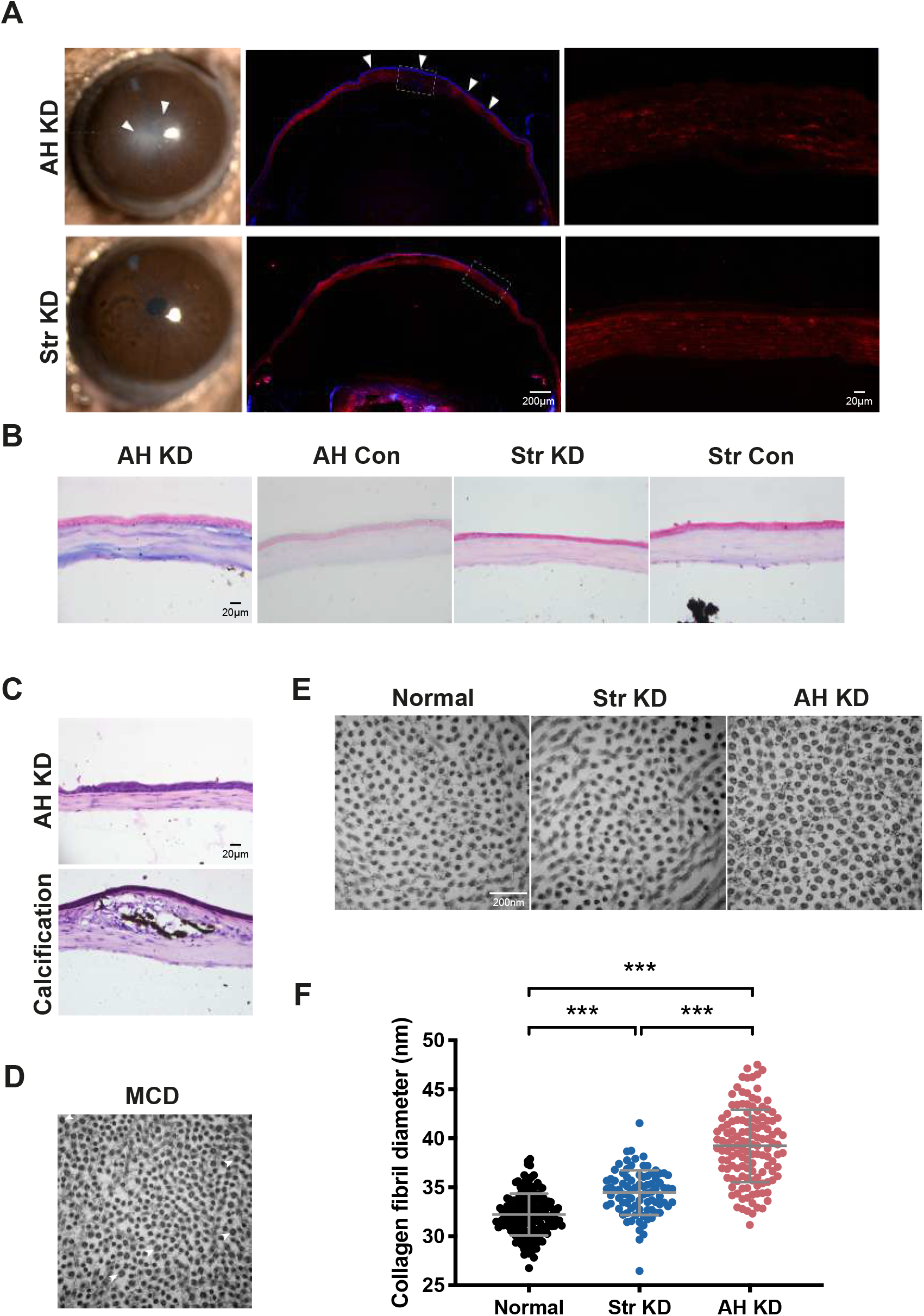
Characterization of *Chst5* knockdown corneas. (*A*) Keratan sulfate staining of the AH KD and Str KD corneas. There was a depletion of KS in the AH KD cornea especially at the central corneal region (white arrowhead, white dash line box), where the AAV9-shRNA plasmids were released in the anterior chamber. No clear lamellar structure existed in the dark area of the AH KD cornea. There was a slightly decline in KS content in some of the Str KD cornea (white dash line box) but the lamellar structure was still intact. (*B*) Alcian blue staining of mice corneas. Blue deposits were spread throughout the full thickness of the AH KD cornea. No such concentrated positive staining could be observed in the AH Con, Str KD, and Str Con groups. (*C*) von Kossa staining for calcification in an AH KD cornea and a positive control cornea. No calcified deposit could be observed in the cornea, indicating the positive alcian blue was not caused by calcification. For positive calcification staining, brown-black calcium deposits were aggregated in the anterior stroma. (*D*) Transmission electron microscopy of MCD patient’s corneal stroma. Abnormally large collagen fibrils could be spotted (white arrowhead). (*E*) Transmission electron microscopy of mice corneal stroma. (*F*) Quantification of corneal stromal collagen fibril diameter. AH KD mouse had significant larger collagen fibrils than the other two groups (*P* < 0.01, One-Way ANOVA, LSD for post hoc tests).

### Overexpression of CHST6 R50H in the anterior chamber induced MCD-like phenotype

CHST6 has several key domains, including a sulfotransferase domain spanning amino acid residue 42 to 356, and two sulfate donor (PAPS) binding domains (amino acid 49-55 and 202-210) (7). To examine whether intracameral overexpression of Chst5 with compromised enzyme function would induce MCD phenotype, we selected a *CHST6* point mutation identified from our patient that resides within one of the PAPS binding domains (Fig 5A-C). The patient showed bilateral MCD phenotype and OCT indicated a full-thickness abnormality in her corneas (Fig 5A). The patient carried a homozygous CHST6 R50H mutation and both of the patient’s parents carried a heterozygous mutation at the same point (Fig 5B). The R50H was conserved among human CHST6, CHST5 and mouse Chst5 (Fig 5D). Overexpression of the R50H Chst5 by intracameral injection-induced MCD-like stromal opacification similar with Chst5 AH KD mice (Fig 5D), with positive alcian blue staining diffused over the corneal stroma (Fig 5E). Again, no calcification staining was observed (Fig 5E). In summary, both intracamerally knockdown of endogenous Chst5 and overexpression of Chst5 mutation could lead to MCD-like phenotype in the cornea, suggesting a potential role of the endothelium in MCD development.

**Fig. 5.**
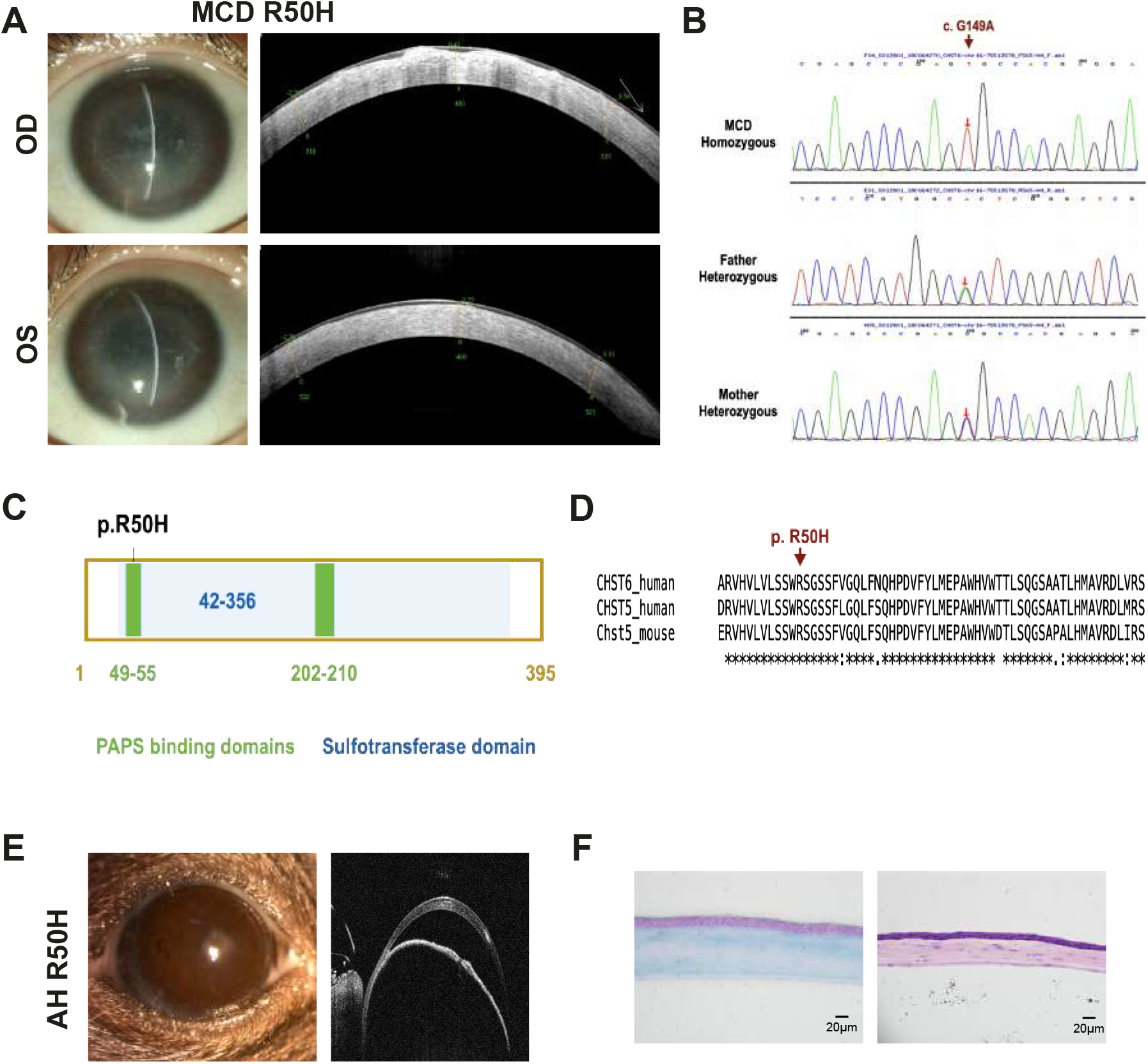
Construction of the *CHST6* mutant overexpression model. (*A*) Eye manifestations of a patient with bilateral MCD presented. (*B*) *CHST6* mutation point status in the pedigree. Exome sequencing identified the patient carrying a homozygous point mutation (c.G148A, p. R50H) in *CHST6*. The patient’s parents carrying heterozygous mutation at the same point and displayed no MCD phenotype. (*C*) Schematic diagram of the CHST6 protein structure. Two PAPS (3’-phospho-5’-adenylyl sulfate) domains are residing at amino acid positions 49-55 and 202-210 (Marked green). The mutation point R50 was inside the first PAPS domain. (*D*) Conservation analysis of the amino acid sequence around the mutation point. The amino acid arginine at CHST6 position 50 is conserved among human CHST6, human CHST5, and mouse Chst5 proteins. (*E*) Eye phenotypes of the mouse with intracameral injection of *Chst5* R50H. White deposit area could be observed in the central corneal region and OCT indicated the opacification involved the full thickness of the cornea. (*F*) Alcian blue and calcification staining of the cornea in (*E*). Positive alcian blue staining spreading at the anterior and posterior stroma. No calcification was detected.

## Discussion

This study retrospectively reviewed the MCD recurrence status in the past 30 years and found PKP could effectively decrease the recurrent rate of MCD, compared with DALK. Whole cornea transcriptomic analysis indicating the MCD pathogenic gene *CHST6* was enriched in corneal endothelium, and so was the CHST6 protein. Knockdown of endothelial *Chst5* or overexpressing of mutant *Chst5* in endothelium by intracameral injection successfully induced MCD-like corneal phenotype in mice. In all, our study gave evidence that MCD is more related to corneal endothelium and provided a new way to construct MCD disease model.

Our retrospective study on MCD patients who received DALK and PKP in the past 30 years and MCD patients who had PTK in the last 10 years in our institute showed an association between surgical methods and recurrence rate, and PKP had a significantly lower recurrence rate compared with DALK, indicating PKP is more effective in reducing MCD recurrence. To our surprise, PTK displayed a lower recurrence rate and longer survival time. It may because PTK is preferred by patients with mild manifestations. Therefore, PTK was not our major focus for comparison. Another interesting observation was that MCD seems recurred from the edges of the graft, suggesting the deteriorative roles played by the leftover corneas.

Though a significant decline of endothelial Chst5 was detected in the AH KD group, we could not exclude the possibility that AAV9-shRNA could enter the corneal stroma and knockdown the keratocyte Chst5. To ascertain the MCD-like phenotype was induced by the lack of endothelial Chst5, we set an intrastromal injection model and did not observe the MCD-like phenotype by directly knocking down the keratocyte Chst5. A more direct way is to create an endothelium-conditional knockdown mouse by using a corneal endothelium-Cre mouse. However, there is still a lack of robust mouse endothelium specific marker to generate the Cre mice, as endothelium and keratocytes have a very close developmental origin. We will explore the mice model by taking advantage of the mouse corneal single cell sequencing results and will examine the Chst5 conditional knockdown effect in the following studies.

Previous studies on Chst5 knockout mice did not observe corneal opacification but found a lack of high-sulfated KS in the cornea (20). Though no visible deposits in the cornea, the synchrotron x-ray fibre diffraction patterns produced by homozygous Chst5-null mice cornea were different from wild-type or heterozygous mice (20). The reason for the absence of corneal opacification in Chst5-null mice was concluded as thin stroma in the mice accompanies by low levels of light scattering (20), and the mouse cornea is predominantly under-sulphated, causing the immunostaining pattern different from other species (9). Despite all these variations in mouse cornea, we noticed corneal transparency impairment in 11.3% of the AH KD mice. Both slit lamp and OCT confirmed the stromal opacification. The deposit area showed a lack of KS staining and was negative for calcification von Kossa staining, indicating they were more resemble MCD deposits but not calcium deposits. The different observations on Chst5 knockdown corneas between the above-mentioned studies might because of the different systems used to generate Chst5-deficient mice, and the following examinations were varied as well. Though not all the examinations such as x-ray diffraction were applied in our study, the full set of controls we used gained strength to our results. There were limitations of our mouse model. MCD need time to develop and most of our mice were still in the very early stage of the disease. TEM only observed abnormal collagen fibrils with a larger diameter in the posterior cornea but did not found vesicles filled with granular materials as was demonstrated in MCD patients (21), or extracellular materials between and among lamellae (22). No mouse with MCD deposits all over the cornea was constructed neither. As only around 1 µL liquid could be injected into the anterior chamber each time, more MCD-like mouse might be constructed by increasing the concentration of the injected AAV9-plasmid and by extending the development time.

It is still not very well understood that how the endothelial Chst5 regulated corneal stromal KS sulphation. Currently, the most vital roles of the corneal endothelium are the barrier function and the pump function. Stroma sulphated KS are highly negatively charged and hydrophilic (23), thus tend to swell. The endothelium is required to dehydrate the stroma and to transport nutrient from the aqueous humour to the stroma (24). However, the mechanism of large molecules transportation from endothelium to the stroma is still not clear. The collagen-rich Descemet’s membrane is up to 10 µm thick and acting as a barrier during the transportation, if there were any exists. Finally, the core structure of corneal KS is synthesized at the rough endoplasmic reticulum and Golgi apparatus and then processed by a series of enzymes, including cGn6ST (23). Suppose the endothelial cGn6ST could translocate to the stroma, how the allochthonous enzyme processing its substrate needs further investigation as well.

Our study suggested a pivotal role of the endothelium in MCD development and recurrence, therefore PKP seems to be a viable choice for MCD surgery. PTK could increase BCVA for a limited time but is likely to have a recurrence in the short term (15). However, some patients preferred PTK rather than corneal transplantation due to psychological reasons. Careful explanations of the consequences are required for these patients.

### Materials and Methods Ethics and subjects

This study was conducted under the tenets of the Declaration of Helsinki and approved by the Ethics Committee of Qingdao Eye Hospital with informed consent obtained from the participants. The data used in this manuscript has been approved by the Review Board of Qingdao Eye Institute.

Patients diagnosed as MCD and underwent LKP or PKP between 1992 and 2021 or underwent PTK between 2011 and 2021 at Qingdao Eye Hospital of Shandong First Medical University were reviewed retrospectively.

### Single-cell sequencing analysis

Three human whole corneas were acquired and digested as previously described (25). The single-cell suspensions were converted to barcoded scRNA-seq libraries using the Chromium Single Cell 3’ Library and Single Cell 3’ v3 Gel Beads (10× Genomics). Libraries were sequenced on Illumina NovaSeq and mapped to the human genome (build GRCh38) using Cell Ranger software (10x Genomics, version 3.1.0) (26). The feature-barcode matrices were further analyzed by the Seurat R package (version 3.2.2) (27). For each library, cells with detected gene numbers less than 200, and the ratio of mitochondria higher than 0.2 were filtered, and genes with at least one feature count in more than five cells were used for the following analysis. Doublets identified by DoubletFinder (28) were removed. Seurat “NormalizeData” and “ScaleData” were used to normalize and scale the data. 4,000 high variable genes were used to performed linear dimensional reduction. Different libraries were integrated through the identification of cell pairwise correspondences between single cells across datasets. Cells were clustered using the ‘‘FindClusters’’ function at 0.4 resolution. The non-linear dimensional reduction technique, tSNE, was employed to learn the underlying manifold and visualization. Cell types were discriminated by the expression of canonical marker genes for each cluster. Gene expression was visualized on a dimensional reduction plot with “FeaturePlot”.

### AAV9 plasmids design and packaging

AAV9 plasmids were packaged by Shanghai GeneChemCo., Ltd. For the knockdown plasmid, shRNA oligos targeting three positions of mouse *Chst5* (NM_019950) were designed and inserted into the GV478 vector (U6-MCS-CAG-EGFP) with the *BsmBI* restriction site. 5’-CGCTGAGTACTTCGAAATGTC-3’ was designed as the knockdown control sequence.

Both the shRNAs plasmids and a *Chst5* overexpression plasmid used for knockdown efficiency test were transfected into HEK 293T cells. Proteins were collected and the gene knockdown effects were examined with Western blot against the FLAG tag with anti-FLAG M2 antibody (1:2000, F1804, Sigma-Aldrich) and goat anti-mouse IgG-HRP antibody (1:4000, sc-2005, Santa Cruz). A pair of oligos with the highest *Chst5* knockdown efficiency was selected for virus packaging (5’-accggACGCTGCCCTTTGCCAAGATTttcaagagaAATCTTGGCAAAGGGCAGCGTttttt-3’).

For the mutant overexpression plasmid (Chst5 R50H), two primers carrying the mutant nucleotide were designed to generate point mutagenesis (Fm: TCCTCGTGGCACTCGGGCTCGTCCTTCGTGG; Rm: GAGCCCGAGTGCCACGAGGACAGTACCAGC). The full-length *Chst5* carrying R50H was inserted into the GV411 vector (CMV-beta globin-MCS-3Flag-SV40 PolyA) between *BsmBI* and *NheI* restriction sites.

### Construction of the macular corneal dystrophy mice

C57BL/6J mice were randomly allocated for AAV-shRNA injection. Two injection routes were designed: intrastromal and intracameral. 1 µL virus was injected with a 36G NanoFil Syringe (World Precision Instruments Inc, FL, USA).

### Immunofluorescence staining

The mouse eye was embedded in the Tissue-Tek O.C.T. compound (Sakura Finetek, Tokyo, Japan), and sectioned into 7 µm thick sections. The sections were stained with KS (Santa Cruz Biotechnology, sc-73518, 1:10) and fluorescein-conjugated secondary antibody (Goat Anti-Mouse IgG H&L, Alexa Fluor 594, Abcam, ab150116, 1:50). Sections were observed under a confocal microscope (Zeiss LSM 880) or Revolve Fluorescence Microscope (Echo) after counterstaining with DAPI.

### Paraffin sectioning and immunohistochemistry

Corneas from MCD patients were fixed in 4% paraformaldehyde at 4 °C, embedded in paraffin and sectioned into 4 µm thickness sections. Anti-CHST6 (1:200, ab154332, Abcam) diluted in NCM Universal Antibody Diluent (WB500D, New Cell & Molecular, China) were used as primary antibodies.

### Alcian blue staining

The cryosection slides were heated at 60 °C for 15 min and fixed in 4% PFA for 15min, rinsed in 3% acetic acid for 3 min and stained with Alcian Blue solution (pH 2.5, Solarbio, G1560) for 30 min. After washed with distilled water the slides were counterstained with Nuclear Fast Red (Beyotime, C0151) for 5 min, dehydrated in ethanol and mounted.

### von Kossa staining

The cryosection slides were heated at 60 °C for 15 min. The slides were covered with von Kossa solution (Servicebio, G1043) for 40 min with exposure to ultraviolet light. After rinsing with distilled water the slides undergoing the hematoxylin and eosin stain procedure.

### Transmission electron microscopy

Samples were trimmed to less than 1 mm^3^ and were fixed with 2.5% glutaraldehyde in 100 mM Sorensen’s phosphate buffer (pH 7.0) at 4 °C for no less than 4 hrs. Samples were then washed with 0.1 M Sorenson’s phosphate buffer pH 7.2 and had a secondary fixation of 1% osmium tetroxide. Samples were dehydrated sequentially in 70%, 80%, 95% and 100% acetone. Samples were then cleared in propylene oxide and infiltrated sequentially in Epoxy 812 embedding medium (Sigma-Aldrich): propylene oxide mixtures at ratios of 2:1 for 0.5 hr at room temperature, of 1:2 for 1.5 hrs at 37 °C and pure embedding medium for 3 hrs at 37 °C. The samples were embedded in a mould and polymerized at 37 °C for 24 hrs, at 45 °C for 24 hrs, and at 60 °C for 24 hrs. Ultrathin sections of 70 nm were made on an ultramicrotome (Reichert-Jung Ultracut E) and placed on a 200 mesh grid. The ultrathin sections were stained with saturated uranyl acetate and lead citrate for 15 min each. The stained sections were examined under a transmission electron microscope (JEM1200-EX, JEOL).

### RNA extraction, cDNA synthesis and qPCR

Mouse cornea tissue was collected, and the endothelial layer was carefully dissected. Total RNAs were extracted with TransZol Up Plus RNA Kit (TransGen Biotech, Beijing, China) by following the manufacturer’s instructions. RNA concentrations and purities were measured using NanoDrop One (Thermo Fisher Scientific). Complementary DNA (cDNA) was synthesized using the PrimeScript cDNA Synthesis Kit (Takara Bio, Dalian, China) following the manufacturer’s instructions and stored at -20 º C. Premix Taq (TaKaRa, Beijing, China) was used for PCR reaction. *Chst5* (NM_019950) qPCR primers used were: 5’-GCCAGCTCTTCAGCCAACAC-3’, 5’-ATACGTCCATGTCGCATAGGAA-3’.

### Exome sequencing for *CHST6* mutation detection

2 mL peripheral blood was collected from the MCD pedigree with consent forms received. Genomic DNA was extracted from the peripheral blood (QIAamp, Qiagen) and exome sequencing was conducted on Illumina HiSeq X Ten System. Sequencing and data analysis were conducted by MyGenostics Inc. (Beijing, China).

## Data Availability

The authors confirm that the data supporting the findings of this study are available within the article. Single-cell sequencing data of this study are available on request from the corresponding author.

## Acknowledgements

The authors would like to thank Dr. Can Zhao for refining the microinjection method, Ting Liu for providing corneal paraffin sections from MCD patients, Rui Cao and Li Gao for providing human corneal tissues, Dr. Wai Kit Chu for suggestions to this study. This study was supported by Shandong Provincial Natural Science Foundation (ZR2020QH140 to B.N.Z.).

## Notes

### Competing Interest Statement

The authors have declared no competing interest.

### Author Declarations

This study was conducted under the tenets of the Declaration of Helsinki and approved by the Ethics Committee of Qingdao Eye Hospital with informed consent obtained from the participants.

